# Crowdsourced partner services among men who have sex with men living with HIV: A pilot randomized controlled trial in China

**DOI:** 10.1101/2022.12.21.22283711

**Authors:** Xumeng Yan, Joseph D. Tucker, William C. Miller, Weiming Tang

## Abstract

**Background:** To improve the uptake of HIV partner services (HIV PS) among men who have sex with men living with HIV (MLWH) in China, our team used a crowdsourcing approach to develop a tailored intervention package. This study assessed the acceptability, feasibility, and preliminary effectiveness of a crowdsourced HIV PS intervention compared to conventional HIV PS.

**Methods:** The study conducted a pilot two-arm randomized controlled trial (RCT) to compare the proportion of HIV testing among sexual partners of MLWH. Indexes in the control arm received conventional HIV PS using referral cards. Indexes in the intervention arm received a crowdsourced HIV PS intervention which included HIV self-testing kits for secondary distribution (HIVST-SD), digital education materials, and assisted PS via provider/contract referral. The primary outcomes were (1) intervention feasibility (i.e., the frequency of indexes using crowdsourced intervention components), (2) intervention acceptability (i.e., the evaluation of indexes on intervention components using Likert scales), and (3) the preliminary impact of the intervention (i.e., the proportion of partners getting HIV testing within three months of index enrollment). Descriptive analysis was conducted, and Chi-squared tests were used to test whether the proportional differences were significant.

**Results:** A total of 121 MSM newly diagnosed with HIV were enrolled between July 2021 and May 2022 in Guangzhou and Zhuhai, China, with 81 in the intervention arm and 40 in the control arm. The 3-month follow-up rates were 93% (75/81) and 83% (33/40), respectively. The crowdsourced intervention components were feasible, as 31 indexes received and 23 indexes used HIVST-SD, 6 indexes used provider-referral to notify 9 sexual partners, and indexes visited the digital educational materials 2.3 times on average. The intervention components also demonstrated acceptability, with HIVST-SD rated 4.4 out of 5 and the digital educational materials rated 4.1 out of 5. The proportion of partners getting HIV testing among all identified partners was 38% (65/171) in the intervention arm, compared to 27% (24/89) in the control arm. The difference was not statistically significant.

**Conclusion:** The crowdsourced HIV PS intervention components were acceptable and feasible among Chinese MLWH and may improve the proportion of stable partners receiving HIV testing. Further implementation science research is needed to expand PS among key populations in low and middle-income countries.

**Clinical trial registration:** **19-0496**

## Introduction

The HIV epidemic continues to expand among Chinese men who have sex with men (MSM).[1] The national data in 2020 show that around 8% of MSM are living with HIV. [2] HIV testing services need to be optimized to reach the key populations who are unaware of their HIV status early in their infection. HIV partner services (PS) is a recommended approach to deliver focused HIV testing services as part of a comprehensive package of HIV testing and care.[3] Defined as a voluntary process for index people living with HIV (PLWH) to inform their partners of potential exposure to HIV and guide them to HIV testing services, HIV PS can improve HIV case-finding, prevent onward transmission, and link people newly diagnosed with HIV into treatment and care.[4]

China’s AIDS Prevention and Control legislation states that PLWH have a duty to inform sexual partner(s) of the diagnosis in an honest and timely manner.[5] Yet, conventional HIV PS using referral cards have made it difficult for healthcare professionals to encourage newly diagnosed patients to disclose to their sexual partners and have their partners present at HIV testing clinics[6], and the uptake of HIV PS remains low in China[7]. Stigma, privacy concerns, and worries about possible negative consequences discourage MSM PS service uptake.[8,9] In response, there have been several new strategies developed to improve the uptake of HIV PS among Chinese MSM, such as an online anonymous partner notification system,[10] community-based-organization (CBO)-based PS mobilization and HIV rapid testing,[11] and assisted HIV PS that provided the options of HIV self-testing and CBO-assisted anonymous notification.[12]

However, these innovative HIV PS strategies may have certain limitations due to the lack of input from key populations. For example, previous studies develop interventions through top-down processes, which have limited contribution from the key clients even when the intervention implementation essentially relies on cooperation with community-based organizations.[10–12] In comparison, crowdsourcing solicits novel solutions through the collective knowledge of the community,[13] and has the strengths of generating greater innovation, fostering multisectoral collaboration, and empowering key populations.[14] Drawing on the experience of using crowdsourcing to promote HIV testing among Chinese MSM,[15,16] our team organized a crowdsourcing open call and a designathon in 2020 to develop interventions to improve the uptake of HIV PS. Crowdsourcing has a group of individuals solve all or part of a problem, then share solutions with the public. A designathon is a type of crowdsourcing event that gather participants to collaboratively create, design and present their solutions within a limited amount of time.[17]

To assess the acceptability, feasibility, and preliminary effectiveness of the crowdsourced HIV PS intervention package in comparison to conventional HIV PS, this study aims to conduct a pilot randomized controlled trial (RCT) among Chinese MSM. These data will help improve HIV PS service delivery in China among MSM.

## Methods

### Intervention Development

Our SESH (Social Entrepreneurship to Spur Health) team held two-stage crowdsourcing events to develop the HIV PS intervention package. From July 10th to September 15th 2020, we held an open call in Guangzhou and received 53 eligible intervention materials.[8] Exceptional submissions from the open call were used as the resources for the subsequent Designathon, which was held from December 18th to December 20th 2020 in Guangzhou. Among eight teams that each delivered a comprehensive intervention package, one team was recognized as the finalist. Members from the winning team then discussed with local healthcare providers and the SESH team to finalize the intervention package components.

### Inclusion Criteria and Study Design

This study used a pilot RCT to evaluate the effect of crowdsourced HIV PS interventions on the uptake of HIV testing among sexual partners of MSM living with HIV (MLWH). Three HIV voluntary counseling and testing (VCT) clinics were selected as recruitment sites; they were based at Guangzhou CDC, an MSM CBO in Guangzhou, and an MSM CBO in Zhuhai, respectively. The Guangzhou CDC provides free HIV confirmatory testing, and the two local CBO sites provide free, community-based HIV testing services for MSM. All the MSM identified in Guangzhou/Zhuhai were referred to these sites for further services. People newly identified with HIV at the HIV VCT clinics would be eligible if they met the following criteria: born male, aged 18 years or older, ever had sex with males, and self-reported having any sexual partners in the last 6 months. Participants were required to provide informed consent for study participation.

### Randomization

Participants were randomly assigned to the intervention group or the control group in a 2:1 ratio. The randomization sequence was created using Microsoft Excel 2022 using random block sizes of three, by a research assistant with no involvement in the HIV VCT clinics. Given the nature of the intervention, there was no blinding to participants or study staff.

### Study Procedures

Upon enrollment, trained investigators at the HIV VCT clinics disseminated baseline surveys to collect information on sociodemographic and behavioral characteristics, sex and HIV testing history, information on each sexual partner in the last 6 months, and preferred referral method for each partner. Participants could choose from self-referral, provider-referral, contract-referral, lost contact, or refusal to disclose. Then, participants were provided crowdsourced HIV partner services or conventional passive HIV partner services according to their allocated groups. They were encouraged by the investigator to disclose their HIV status to sexual partners and get their partners tested. Indexes were contacted for follow-up surveys three months after enrollment, to collect data on whether their partners were notified of the HIV exposure risk, the actual referral method, partner uptake of HIV testing, and the HIV testing results. (Sfigure 1) Indexes who self-reported their partners as testing positive would be contacted by investigators to provide contact information of these partners with informed consent. Men were enrolled from July 13^th^ 2021 to May 26^th^ 2022. This trial was registered on ClinicalTrials.gov (19-0496).

**Figure 1.**
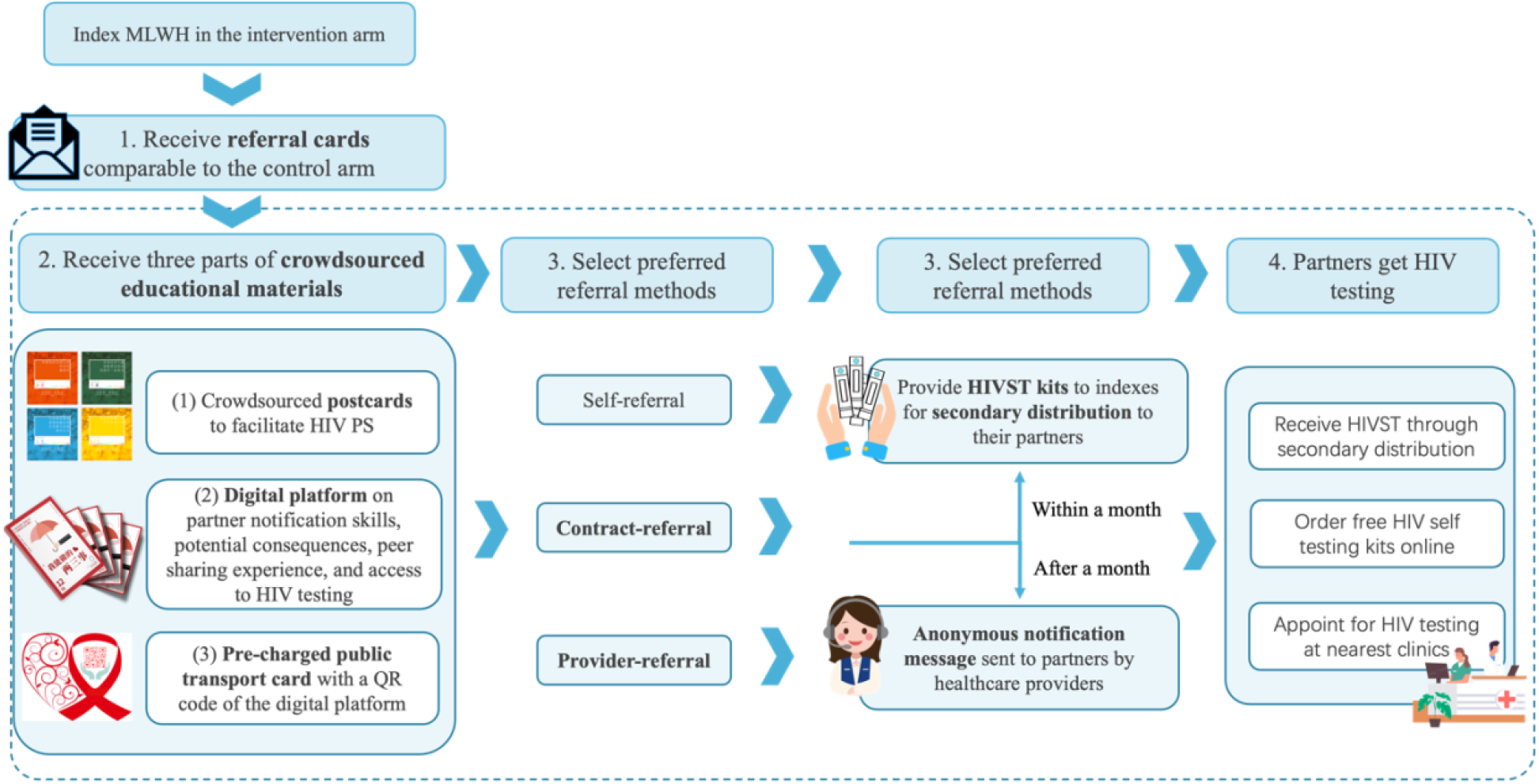
Intervention components and procedures for indexes in the intervention arm

### PS delivery

In the control group, participants were offered passive referral using referral cards. We used this referral method because it is currently the main method routinely used in China.[18] The referral cards had three QR codes on them: one for appointing onsite HIV testing services at the HIV VCT clinics nearby, one for applying for free HIV self-testing (HIVST) kits online (mailed to the given addresses), and one for sexual partners to upload a picture or a snapshot of their HIV testing results. Sexual partners who uploaded their testing results were offered $3.5 as an incentive, and their returned data would be linked to their indexes through a unique ID assigned to the indexes at enrollment. Indexes were offered $7.5 as an incentive for each identifiable sexual partner whom they successfully motivated to upload their testing results.

In the intervention group, participants received the same referral cards, and the same incentives were provided for the participants and their sexual partners. Aside from that, they were provided with crowdsourced HIV partner services that integrated three parts: (a) educational materials for HIV PS promotion purposes, (b) secondary distribution of HIVST kits (HIVST-SD) to sexual partners, and (c) options of provider referral or contract referral using anonymous notification messages. (Figure 1)

The educational materials included (a.1) postcards from previous crowdsourcing events to facilitate HIV PS, and (a.2) a digital platform embedded in the WeChat application that offered information on what is HIV PS, why HIV PS is important, partner notification skills by partner types, and potential risks or adverse consequences. The digital platform also included a section on peer sharing, where MLWH shared their experience of successfully disclosing their HIV status to their sexual partners in text or video forms. The digital platform did not track any identifying information of the users and encouraged the use of aliases to protect user privacy. The QR code of the digital platform was printed on (a.3) a public transport card pre-charged with $3 for portable usage and easy access.

As participants were required to select a preferred referral method for each of their self-reported partners, investigators provided participants who selected self-referral with corresponding numbers of HIVST kits and instructed the participants to distribute the HIVST kits to their sexual partners. When participants selected provider-referral or contract-referral, they were required to leave the contact information of their sexual partners (phone numbers, emails, or social media accounts such as Blued or WeChat were all acceptable). Participants who selected contract-referral were given one month to finish self-referral. Provider-referral would be initiated if they did not disclose their HIV status to their sexual partners within one month. When initiating provider-referral, investigators would contact the sexual partners and send the following message: “Hello. This message is generated by XXX HIV VCT clinic. Have you been exposed to unprotected sex without getting tested for HIV? We care about your health and would encourage you to get HIV testing at your convenience. You can appoint HIV testing services at your nearest clinic or click the <u>link A</u> to apply for free HIVST kits for at-home testing. Click the <u>link B</u> to upload a photograph of your testing results, and you will be compensated $3.5. You can rest assured that your privacy is protected by us. If you have any concerns regarding this message, please reply directly to start counseling.” The notification message was anonymous as it did not reveal any index information.

### Outcome Measurements

The primary outcomes were (1) the feasibility of the study and the feasibility of the intervention, (2) the acceptability of the intervention, (3) the preliminary impact of the intervention, which was the proportion of partners getting HIV testing within 3 months of the index participant’s enrollment. The feasibility of the study was measured in the ability to enroll newly identified MLWH as planned within a 12-month period and the 3-month follow-up rate of the indexes and their partners. The feasibility of the intervention was measured by the frequency of the indexes and their partners in the intervention arm receiving and using the crowdsourced intervention components. The acceptability of the crowdsourced intervention was measured at follow-up surveys of indexes and their partners in the intervention arm, who would be asked to evaluate the interventions. The preliminary impact of the intervention was measured through the proportion of indexes who successfully had one or more partners tested for HIV within 3-month follow-up and the proportion of sexual partners who received HIV testing. Whether a sexual partner received HIV testing was based on the indexes’ self-reported information at 3-month follow-up surveys. We also reported the proportion of sexual partners who uploaded HIV testing results and who uploaded HIV-positive results.

The secondary outcomes included (1) the proportion of sexual partners who were notified by indexes, (2) the proportion of sexual partners who tested positive among tested partners, (3) the mean number of partners notified per index, (4) the mean number of partners tested per index, and (5) the mean number of case-finding (i.e., partners tested positive) per index. All the outcomes were stratified by partner types.

A stable male sexual partner was defined as a male with whom the index had a sexual relationship lasting more than three months in the past six months and a casual male sexual partner was defined as a man with whom the index had occasional sex in the past six months. A female sexual partner was defined as a woman having any sexual relationship with the index case in the past six months. We further asked detailed questions about the index’s definition of their sexual relationship categorized by the level of commitment, including spouse/long-term stable partners, boyfriends/girlfriends, dating relationships, friends with benefits, booty calls, and one-night stands.

### Statistical Analysis

Descriptive analyses were conducted to report the demographic and behavioral characteristics of the indexes, and to assess the feasibility and acceptability of the study and the crowdsourced interventions. In addition, we compared the HIV testing cascade in the intervention group to that in the control group, stratified by sexual partner types. The HIV testing cascade included: (a) the number of self-reported sexual partners, (b) the number and proportion of sexual partners who were notified of the index’s HIV status and their proportion among those who were reported, (c) the number of sexual partners who received HIV testing and their proportion among those who were notified, and (d) the number of sexual partners who tested positive and their proportion among those who got tested. These results were based on index self-report follow-up surveys and were compared using a Chi-squared test. All analysis was done using R version 4.2.1.

## Results

From July 13^th^ 2021 to May 26^th^ 2022, 263 MSM newly diagnosed with HIV were screened for eligibility, 58 were not eligible. An additional 84 did not provide informed consent and were excluded from the study. A total of 121 eligible MSM were recruited, with 81 assigned to the intervention group and 40 assigned to the control group. At the three-month follow-up, 75 of 81 (93%) participants from the intervention group and 33 of 40 (83%) participants from the control group were successfully contacted. (Figure 2)

**Figure 2.**
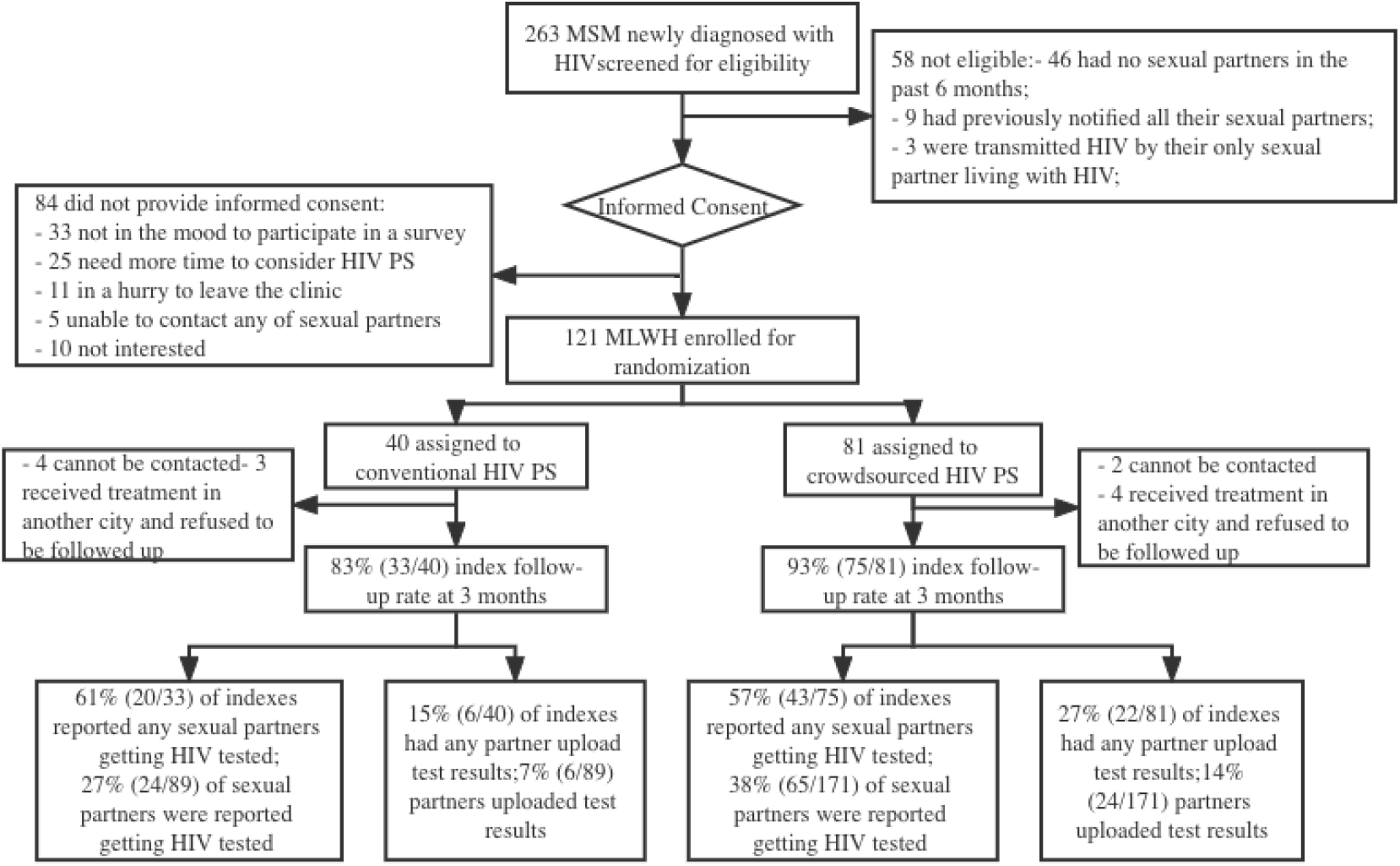
Summary of study enrollment, follow-up, and key outcomes among MSM living with HIV in China, n = 263, 2021-2022

### Descriptive statistics of indexes at the baseline survey

Of 121 participants, the mean age was 30.2 years (standard deviation (SD): 9.24), 82% were single, and more than half of the participants had a college degree or above, and 75% self-reported their sexual orientation as gay. Nearly 80% of participants ever disclosed their sexual orientation to others. Regarding sexual behaviors, 56% reported no condom usage in the most recent sex, 45% reported being receptive in sex roles, 47% reported ever using Rush or other drugs during sex, and 22% reported a previous diagnosis of STI other than HIV. Participant characteristics were similar in the intervention and the control groups. (Table 1)

**Table 1.** Descriptive characteristics of 121 indexes at baseline, 2021-2022, Guangdong, China.

| Variables | Group | Total<br>(n=121, %) | Treatment<br>(n=81) | Control<br>(n=40) |
| --- | --- | --- | --- | --- |
| Average age |  | 30.2<br>(SD: 9.24) | 29.8<br>(SD: 9.09) | 31.0<br>(SD: 9.61) |
| Site | Guangzhou | 54 (45%) | 35 (48%) | 19 (47%) |
|  | Zhuhai | 67 (55%) | 46 (52%) | 21 (53%) |
| Marital status | Married or engaged | 12 (10%) | 8 (10%) | 4 (10%) |
|  | Single | 99 (82%) | 67 (83%) | 26 (80%) |
|  | Divorced or widowed | 10 (8%) | 6 (7%) | 4 (10%) |
| Education level | Middle school or below | 26 (22%) | 14 (17%) | 12 (30%) |
|  | High school | 27 (22%) | 21 (26%) | 6 (15%) |
|  | College/Dazhuan | 66 (54%) | 44 (55%) | 22 (55%) |
|  | Graduate or above | 2 (2%) | 2 (2%) | 0 (0%) |
| Annual income (in USD) | ≤3000 | 22 (18%) | 16 (20%) | 6 (15%) |
|  | 3001-6000 | 8 (7%) | 6 (7%) | 2 (5%) |
|  | 6001-10000 | 33 (27%) | 21 (26%) | 12 (30%) |
|  | 10001-16000 | 29 (24%) | 21 (26%) | 8 (20%) |
|  | >16000 | 29 (24%) | 17 (21%) | 12 (30%) |
| Sexual orientation | Gay | 91 (75%) | 56 (69%) | 35 (87%) |
|  | Straight | 6 (5%) | 4 (5%) | 2 (5%) |
|  | Bisexual | 21 (17%) | 18 (22%) | 3 (8%) |
|  | Other/Prefer not to answer | 3 (3%) | 3 (4%) | 0 (0%) |
| Sexual orientation disclosure | Yes | 95 (79%) | 64 (79%) | 31 (77%) |
|  | No | 26 (21%) | 17 (21%) | 9 (23%) |
| Previous STI diagnosis (HIV excluded) | Yes | 27 (22%) | 11 (14%) | 16 (40%) |
|  | No | 94 (78%) | 70 (86%) | 24 (60%) |
| Had sex with males within 6 months | Yes | 117 (97%) | 79 (98%) | 38 (95%) |
|  | No | 4 (3%) | 2 (2%) | 2 (5%) |
| Condom frequency with males within 6 months | Never | 29 (25%) | 20 (25%) | 9 (24%) |
|  | Seldom | 27 (23%) | 17 (22%) | 10 (26%) |
|  | Often | 36 (31%) | 26 (33%) | 10 (26%) |
|  | Every time | 25 (21%) | 16 (20%) | 9 (24%) |
| Condom usage in most recent sex | Yes | 53 (44%) | 32 (40%) | 21 (53%) |
|  | No | 68 (56%) | 49 (60%) | 19 (47%) |
| Sex role | Insertive | 26 (22%) | 19 (24%) | 7 (17%) |
|  | Receptive | 55 (45%) | 35 (43%) | 20 (50%) |
|  | Both | 40 (33%) | 27 (33%) | 13 (33%) |
| Used Rush & other drugs during sex | Yes | 47 (47%) | 38 (47%) | 19 (47%) |
|  | No | 64 (53%) | 43 (53%) | 21 (53%) |
| Reported number of stable male partners | 0 | 53 (44%) | 34 (42%) | 19 (48%) |
|  | 1 | 56 (46%) | 41 (51%) | 15 (38%) |
|  | 2 | 11 (9%) | 6 (7%) | 5 (12%) |
|  | 3 or more | 1 (1%) | 0 (0%) | 1 (2%) |
|  | 0 | 42 (35%) | 32 (40%) | 10 (25%) |
| Reported number of casual male partners | 1 | 42 (35%) | 26 (32%) | 16 (40%) |
|  | 2 | 15 (12%) | 8 (10%) | 7 (18%) |
|  | 3 or more | 22 (18%) | 15 (18%) | 7 (17%) |
| Reported number of female partners | 0 | 111 (92%) | 74 (91%) | 37 (93%) |
|  | 1 or more | 10 (8%) | 7 (9%) | 3 (7%) |

### Feasibility & acceptability

The pilot study was feasible in recruitment and follow-up with indexes as it enrolled newly identified MLWH between July 13^th^ 2021 and May 26^th^ 2022, and had a total follow-up rate of 89%, (83% in the control arm and 93% in the intervention arm, respectively). Yet, the rate of sexual partners uploading their HIV testing results was quite low. Of 89 identified partners in the control arm, 24 (27%) partners were reported as having tested for HIV by their index, while only 6 (7%, 6/89) partners uploaded their testing results. Of 171 identified partners in the intervention arm, 65 (38%) partners were reported as having tested for HIV by their index, while 24 (14%, 24/171) partners uploaded their testing results. To verify the self-reported results, local investigators checked that the index self-reported information of partners testing HIV positive was accordant with HIV registration records, which allowed us to use the self-reported notification results as a proxy measurement to reflect the actual HIV PS uptake.

The crowdsourced interventions demonstrated feasibility: (1) the digital platform that contained educational materials had 276 hits from 121 visitors during the study period, an average of 2.3 hits per visitor; (2) 31 indexes in the intervention arm obtained HIVST-SD at baseline, among whom 23 indexes reported successful secondary distribution to their sexual partners at follow-up surveys; (3) 9 sexual partners of 6 indexes in the intervention arm were successfully notified using provider-referral.

The crowdsourced interventions were also acceptable. Indexed were asked to evaluate whether they were happy to receive the crowdsourced components at the follow-up survey using a 5-point Likert scale, where 1 means strongly disagree and 5 means strongly agree. Among 57 indexes in the intervention arm who completed the evaluation questions, the average rating score of crowdsourced postcards was 3.7 out of 5, and the scores of the digital platform and the transport card that had a QR code for the digital platform were 4.1/5, and 3.6/5. Among 31 indexes that received HIVST-SD, the average rating score was 4.4/5. Among these components, the HIVST-SD and digital platform showed the highest acceptability.

### Self-reported partner notification and testing rates for indexes

Of 75 indexes in the intervention group who completed follow-up surveys, 47 (62.7%) notified at least one of their reported sexual partners, including 25 (33.3%) who notified all their partners and 22 (29.3%) who notified part of their sexual partners. Of 33 indexes followed up in the control group, 20 (60.6%) notified at least one of their reported sexual partners, including 9 (27.2%) who notified all their partners and 11 (33.3%) who notified part of their sexual partners. The differences between the intervention group and the control group were not statistically significant. (Stable 1)

### HIV testing cascades by type of sexual partner

The total number of identified sexual partners of any type was 171 in the intervention group, including 53 stable male partners, 110 casual male partners, and 8 female partners. The total number of identified partners was 89 in the control group, including 27 stable male partners, 58 casual male partners, and 4 female partners. The number of sexual partners identified per index was 2.1 in the intervention arm and 2.2 in the control arm.

For the primary outcome, based on the 3-month follow-up of index self-reported outcomes, the proportion of sexual partners who received HIV testing among all identified partners was 38% (65/171) in the intervention group and 27% (24/89) in the control group. The proportions of partners getting tested among stable male partners were 68% (36/53) in the intervention arm and 59% (16/27) in the control arm; the proportions of partners getting tested among casual male partners were 22% (24/110) and 10% (6/58) in the two groups, respectively.

The HIV testing cascade showed that the rates of being notified were all higher in the intervention group than in the control group (35% vs. 28% for total partners, 74% vs. 59% for stable male partners, and 13% vs. 12% for casual male partners). Among sexual partners who were notified, the percentage of those who had HIV testing was higher but did not differ significantly between the intervention group (110%) and the control group (96%). Among partners getting HIV testing, the rates of testing positive were higher in the intervention group (23% vs. 13% for total partners, 39% vs. 13% for stable male partners, and 4% vs. 0% for casual male partners). (Figure 3) None of the proportion differences were statistically significant. The number of sexual partners notified/tested for HIV/tested positive per followed-up index was 0.8/0.9/0.2 in the intervention arm, and 0.8/0.7/0.1 in the control arm. The intervention group had slightly higher self-reported partner testing and case-finding numbers per index. It is noted that tested sexual partners outnumbered notified sexual partners in the intervention group because our interventions did not require indexes to disclose their HIV status as a necessary procedure to get their sexual partners tested.

**Figure 3.**
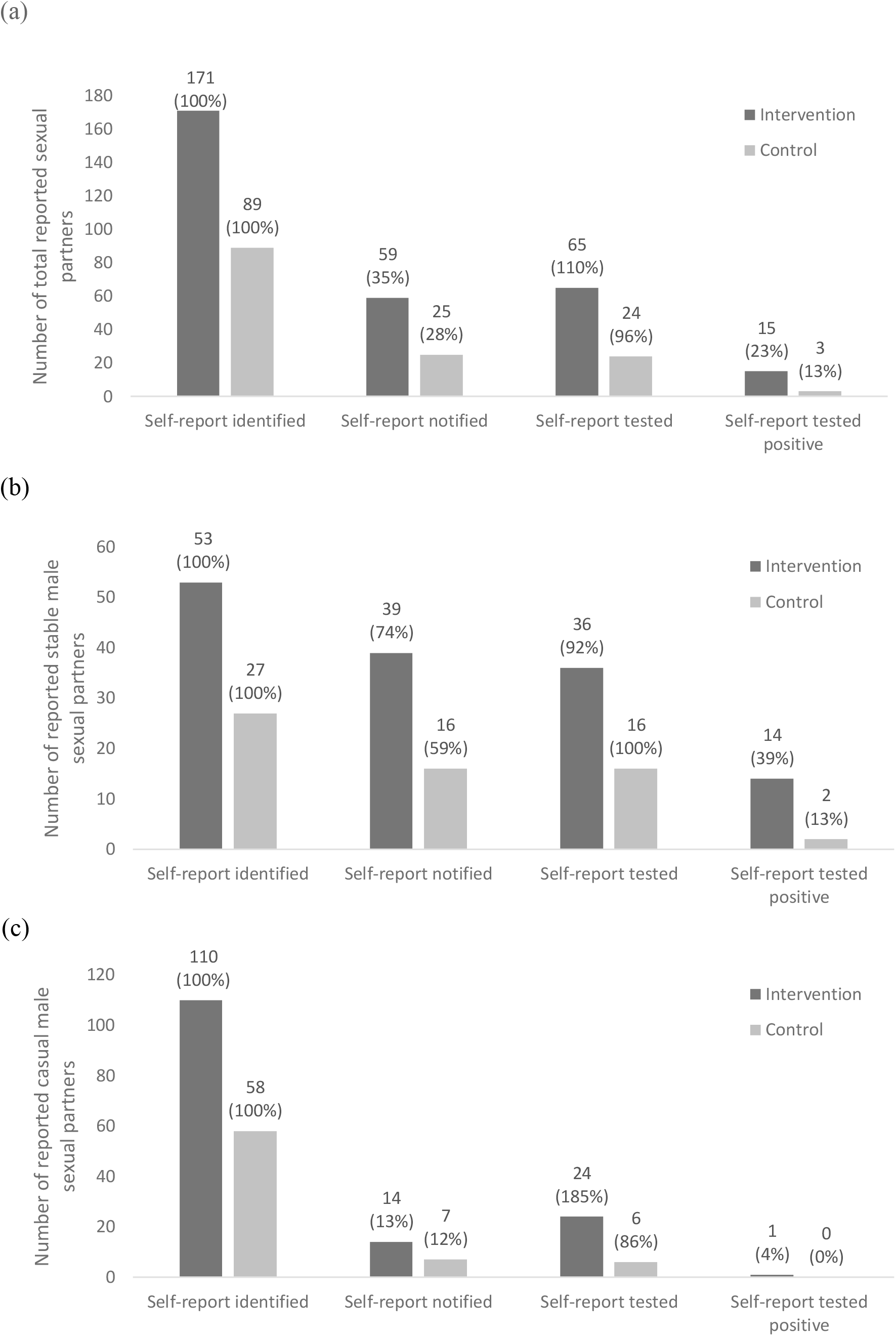
HIV testing cascades in the intervention group and the control group, by type of sexual partners. (a) HIV testing cascade for all sexual partners; (b) HIV testing cascade for stable male sexual partners; (c) HIV testing cascade for casual male sexual partners. The percentage in the parenthesis for each bar was obtained by comparing it with the number in the left adjacent bar in the cascade.

At baseline, indexes were required to indicate their preferred referral methods. In the intervention group, 108 of 171 (64%) partners were identified as contactable partners as the indexes did not lose contact with them or refuse to disclose their HIV status; and 55 of 89 (62%) partners in the control group were contactable partners. At 3-month follow-up, 59 of 108 (55%) contactable partners were notified in the intervention arm, including 50 using self-referral and 9 using provider-referral, while 25 of 55 (45%) contactable partners were notified in the control arm, all using self-referral. (Table 2)

**Table 2.** Comparison of baseline and follow-up referral methods for contactable partners

|  | Preferred methods at baseline |  | Reported methods at follow-up |  |
| --- | --- | --- | --- | --- |
| Referral methods | Intervention (N index=81) | Control (N index=40) | Intervention (N index=75) | Control (N index=33) |
| Self-referral | 84 (49%) | 52 (58%) | 50 (85%) | 25 (100%) |
| Provider-referral | 14 (8%) | 3 (3%) | 9 (15%) | 0 (0%) |
| Contract-referral | 10 (6%) | 0 (0%) | 0 (0%) | 0 (0%) |
| Refuse disclosure | 5 (3%) | 1 (1%) | - | - |
| Lost contact | 58 (34%) | 33 (37%) | - | - |
| **Contactable total** | **108 (64%)** | **55 (62%)** |  |  |
| **Total** | **171 (100%)** | **89 (100%)** | **59 (100%)** | **25 (100%)** |

In both arms, the proportion of partners uploading testing results was low, yet it was slightly higher for the intervention group. Of 24 (14%) partners in the intervention group who returned testing results, 15 used HIVST (including nine who used HIVST-SD), and three tested positive. Only 6 (7%) partners in the control group returned testing results, among whom four used HIVST and one tested positive. Most of these partners who returned testing results were spouses/stable partners, including 19 (79%, 19/24) in the intervention arm and 6 (100%, 6/6) in the control arm.

### Positive/negative consequences reported

Five indexes (four in the intervention arm and one index in the control arm) reported psychological stress getting relieved after HIV disclosure. Three indexes reported becoming more intimate with their partners and three indexes reported receiving support from their partners. One index in the intervention arm reported getting divorced from his female partner as well as suffering violence other than intimate relationship violence. The index was contacted by local investigators and referred to free counseling support.

## Discussion

The pilot results provide preliminary evidence suggesting that a crowdsourced HIV PS intervention was acceptable and feasible among Chinese MLWH and their sexual partners. While the rates of indexes notifying any sexual partner and getting any partner tested were similar in two arms, indexes in the intervention arm had higher rates of notifying all partners and higher rates of getting all partners tested. The HIV testing cascades also showed that sexual partners linked to indexes in the intervention group had higher proportions of getting HIV testing among all identified partners (38% vs 27%). Among contactable partners, the percentage of partners getting HIV testing was also higher (55%) in the intervention group than in the control group (45%). This study expands the literature on HIV PS by using a novel community-based crowdsourcing intervention package and focusing on MSM diagnosed with HIV in resource-limited settings.

Our crowdsourced HIV PS intervention package demonstrated preliminary impact in improving the uptake of partner HIV testing and case-finding. The proportions of all/stable/casual partners getting tested in the intervention arm were 38%, 68%, and 22%, while the corresponding proportions in the control arm were 27%, 59%, and 10%. Additionally, the intervention package had higher rates of case-finding compared to conventional referral cards. Twenty-three percent of tested partners were tested positive in the intervention arm, versus 13% in the control arm. The case-finding rate was even higher for stable male partners, with 39% in the intervention arm and 13% in the control arm. Meanwhile, the crowdsourced intervention brought substantial psychological benefits, as the indexes reported positive outcomes associated with the use of HIV PS, including reducing psychological stress and improved relationship with partners. This was consistent with previous studies that most partners would express psychological support for the index after HIV PS. [7] Our pilot study provided evidence for further implementation of the crowdsourced HIV PS among Chinese MLWH in a full RCT.

Among the crowdsourced components, integrating HIVST-SD into HIV PS was the most promising intervention. Our results indicated that a large proportion (74%) of indexes who received the HIVST-SD intervention at baseline reported successful secondary distribution of HIVST to their sexual partners, and 9 of 24 partners who uploaded testing results reported receiving HIVST-SD from the indexes. Previous RCTs have evidence that HIVST-SD was effective in promoting peer testing and partner testing mostly among women and their male partners, such as antenatal and postpartum women, female sexual workers and their male partners in Kenya, [19,20] young women and their partners in South Africa. [21] Several studies have reported the impact of HIVST-SD on promoting first-time HIV testing and testing frequency among MSM and their peers and/or sexual partners. [22,23] Our study suggests that HIVST-SD may be incorporated into HIV PS services for MLWH as well. In the next step, we plan to optimize the integration of HIVST-SD into HIV PS. For example, previous evidence has mentioned that HIVST-SD can be delivered via social media-based digital networks, [24–26] and that machine learning models can be applied to improve the effect of HIVST-SD. [27]

Casual male partners remain a hard-to-reach population for HIV PS, but we found that getting partners tested without disclosure and assisted PS with anonymous notification may be useful for casual partner HIV testing. Indexes in the intervention arm reported more casual male partners getting HIV testing than receiving formal partner notification. This is consistent with our previous finding in a cross-sectional survey and qualitative analysis of the crowdsourcing open call that getting partners tested without HIV disclosure may be a strategic way for indexes to care for their sexual partners and protect their privacy at the same time. [8] The advice of getting partners tested without HIV disclosure was also included in the crowdsourced digital platform, within the section of Casual Partner Notification Skills: “For booty calls, you can suggest regular HIV testing for the safety and health of each other, so that your partner can get tested without HIV disclosure from you.” It is possible that some indexes in the intervention group followed the advice from the digital platform and encouraged their partners to get tested without HIV notification. On the other hand, some indexes chose to use provider referral to contact their casual male partners. Assisted PS has numerous evidence of improving HIV testing uptake, increasing HIV new case identification and improving linkage to treatment and care, [28] especially in resource-limited settings. [29–31] Nevertheless, the uptake of provider-referral/contract-referral in our study was low compared to other studies in different sites. This may be due to that these assisted referral methods were offered as a choice, while most indexes preferred to use self-referral instead of giving the partners’ contact information to providers. The uptake of assisted PS can be improved by addressing key barriers including stigma and discrimination from healthcare providers, [32] indexes needing more time to process HIV-positive status before providing partner notification, and lack of trust in providers. [33] Future studies are needed to tailor these strategies to improve the uptake of HIV PS particularly for casual partners.

Our study has several limitations. First, our primary outcomes were based on self-reported results by the index, which may have reporting bias. Yet, we had investigators contact indexes who reported their sexual partners as testing positive and obtain these partners’ contact information to check with HIV registration records. The index self-reported information of partners testing HIV positive was consistent with HIV registration records. Meanwhile, any reporting bias of partner HIV testing should be similar in both arms, as indexes in both arms were not provided incentives for self-reported partner testing. Second, our sample size did not have sufficient power to test the differences in primary and secondary outcomes. Future studies require full RCTs with a larger sample size to evaluate the actual effectiveness of the crowdsourced interventions. Third, the feasibility and acceptability of the crowdsourced interventions may not be generalizable to MLWH elsewhere, as the finalized interventions were largely co-created by local MLWH, MSM CBOs, and healthcare providers in Guangdong Province, China. However, one strength of the crowdsourcing approach is that it is local wisdom-based and does not look for a one-size-fits-all, standardized solution. Fourth, almost one in every three eligible participants declined to enroll in the study. Most of them reported that they were not ready to participate in a survey or think about partner notification because they still needed time to accept being diagnosed with HIV. Some of them reported a lack of time to enroll in the study at baseline because they would like to initiate same-day ART at an AIDS-designated hospital far away from the HIV VCT clinic. Optimization of the recruitment procedures is needed in future studies, such as allowing a time buffer for indexes to decide, and following up with indexes that declined at first to check whether they changed their mind.

## Conclusion

Overall, the crowdsourced HIV PS interventions that incorporated HIVST-SD, digital educational materials, and assisted PS via provider/contract referral were acceptable and feasible among Chinese MLWH. There was preliminary evidence that the crowdsourced interventions may improve the proportion of partners getting tested for HIV. Future modifications on the intervention components and study procedures are needed to design a full RCT to evaluate the impact of crowdsourced HIV PS interventions.

## Data Availability

All data produced in the present study are available upon reasonable request to the authors

