## Supplemental Figure 1 and Table 1 for "Crowdsourced partner services among men who have sex with men living with HIV: A pilot randomized controlled trial in China"

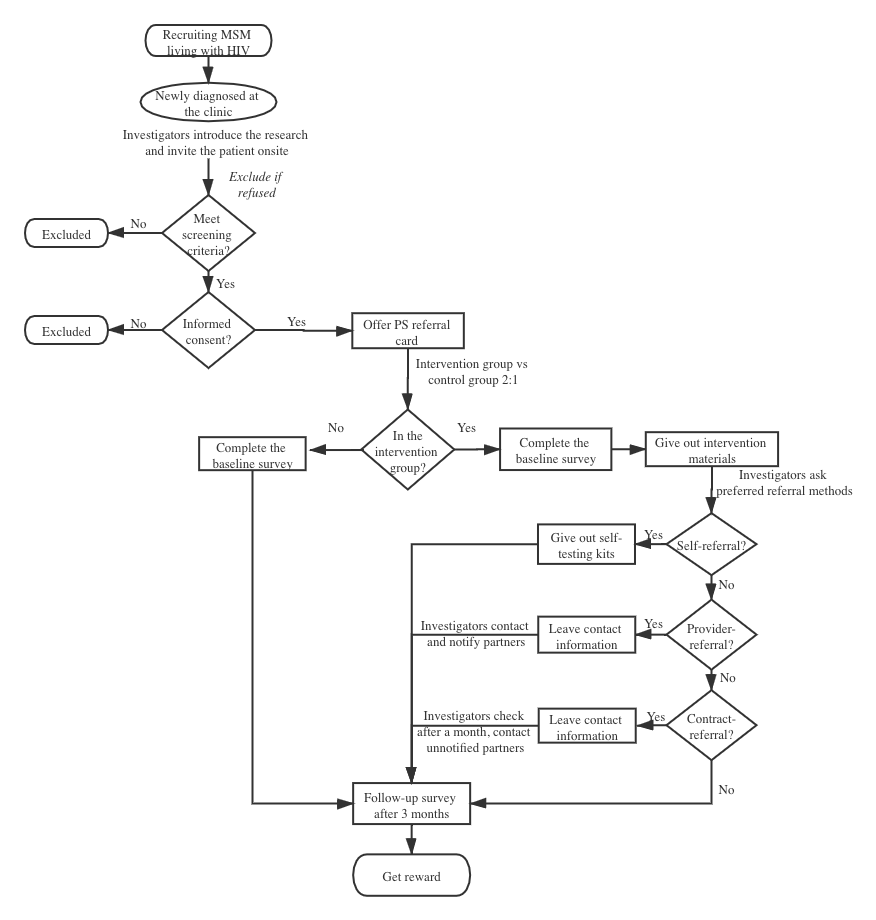

Sfigure 1. Details of the pilot RCT study procedures

Stable 1 Self-reported partner notification and testing rates among 108 followed-up indexes, 2022, Guangdong, China.

| **Number (%)** | **Intervention group (n=75)** | **Control group (n=33)** | **Chi-squared** |
| --- | --- | --- | --- |
| **Self-reported partner notification** |  |  | 0.63 (p=0.73) |
| Notified all sexual partners | 25 (33.3%) | 9 (27.2%) |  |
| Notified some sexual partners | 22 (29.3%) | 11 (33.3%) |  |
| Notified none of sexual partners | 28 (37.3%) | 13 (39.4%) |  |
| **Self-reported partner testing** |  |  | 2.84 (p=0.24) |
| Had all sexual partners tested | 25 (33.3%) | 8 (25.0%) |  |
| Had some sexual partners tested | 18 (24.0%) | 12 (36.4%) |  |
| Had none of sexual partners tested | 32 (42.7%) | 13 (39.4%) |  |
